# Variations of the quality of care during the COVID-19 pandemic affected the mortality rate of non-COVID patients with hip fracture

**DOI:** 10.1101/2021.11.27.21266927

**Authors:** Davide Golinelli, Francesco Sanmarchi, Angelo Capodici, Giorgia Gribaudo, Mattia Altini, Simona Rosa, Francesco Esposito, Maria Pia Fantini, Jacopo Lenzi

**Affiliations:** Department of Biomedical and Neuromotor Sciences (DIBINEM), Alma Mater Studiorum – University of Bologna. Via San Giacomo 12, 40126, Bologna, Italy; Healthcare Administration, AUSL Romagna, Ravenna, Italy

**Keywords:** COVID-19, Hip Fracture, Healthcare Quality, Mortality, Surgery, Pandemic

## Abstract

**Introduction:** As COVID-19 roared through the world, governments worldwide enforced containment measures that affected various treatment pathways, including those for hip fracture (HF). This study aimed to measure process and outcome indicators related to the quality of care provided to non-COVID-19 elderly patients affected by HF in Emilia-Romagna, a region of Italy severely hit by the pandemic.

**Methods:** We collected the hospital discharge records of all patients admitted to the hospitals of Emilia-Romagna with a diagnosis of HF from January to May in the years 2019/2020. We analyzed surgery rate, surgery timeliness, length of hospital stay, timely rehabilitation, and 30-day mortality for each HF patient. We evaluated monthly data (2020 vs. 2019) with the chi-square and t-test, where appropriate. Logistic regression was used to investigate the differences in 30-day mortality.

**Results:** Our study included 5379 patients with HF. In April and May 2020, there was a significant increase in the proportion of HF patients that did not undergo timely surgery. In March 2020, we found a significant increase in mortality (OR = 2.22). Female sex (OR = 0.52), age ≥90 years (OR = 4.33), surgery after 48 hours (OR = 3.08) and not receiving surgery (OR = 6.19) were significantly associated with increased mortality. After adjusting for the aforementioned factors, patients hospitalized in March 2020 still suffered higher mortality (OR = 2.21).

**Conclusions:** Our results show a reduction in the overall quality of care provided to non-COVID-19 elderly patients affected by HF. The mortality rate of patients with HF increased significantly in March 2020. Patients’ characteristics and variations in processes of care partially explained this increase. Our analysis reveals the importance of including process and outcomes indicators, for both acute and post-acute care management issues, in emergency preparedness plans, to monitor healthcare systems’ capacities and capabilities.

## INTRODUCTION

Timely detection, intervention, and rehabilitation are key factors for the successful management of patients who suffer from hip fractures (HFs) [1-5]. Clinical guidelines recommend immediate surgical repair of HF following hospital admission [6-8]; typical hospital stay lasts a few days and thereafter the patient is transferred for rehabilitation [9]. To improve functional outcomes and reduce mortality, international guidelines recommend performing surgery within 48 hours of hospital admission, or as soon as the patient is medically stable, avoiding a delay in surgery, ensuring early mobilization, and providing a post-acute rehabilitation plan [3, 9-11].

In 2020, the COVID-19 pandemic forced governments worldwide to enforce containment measures such as social distancing and home quarantining. These measures had a considerable impact on various treatment pathways [11-13], including those for HF [14]. During the initial phases of the pandemic, elective surgery was halted in many healthcare systems, and only emergencies were treated [15]. In many hospitals, wards were merged, and non-COVID-19 cohorts were created to reduce cross-infection between staff members and patients [16]. Early discharge of HF patients was encouraged to increase the number of available beds, reallocate clinical staff, and ensure patient and staff members’ safety standards.

As COVID-19 roared through the world, treatment pathways for HF felt the blow and musculoskeletal facilities were reorganized as a result, patients were at risk of being left without proper care and were exposed to increased disability and mortality [17,19-21]. Some healthcare systems withstood the impact, maintaining levels of care for non-COVID-19 patients similar to those of the pre-pandemic period. Other systems, such as the Italian one, were caught unprepared and this affected their performance at both the hospital and out-of-hospital level [23].

After the first case of COVID-19 on February 21, 2020, the first wave of the pandemic struck all over northern Italy, including Emilia-Romagna. Italy’s national government decided to promptly implement strict non-pharmaceutical measures. National quarantine was declared, and the entire country was under lockdown from March 9 to May 4; complete freedom of movement was not reintroduced until June 3, 2020 [16]. These restrictive measures affected schools, universities, bars, and restaurants, and determined the disruption of usual healthcare pathways, as healthcare systems had to react to the emergency, suspending non-urgent surgical interventions, outpatient visits, and many primary services.

Healthcare systems’ ability to adapt to the mutating population’s health needs caused by an emergency such as the COVID-19 pandemic can be assessed through several health processes and outcome indicators [24]. Accordingly, it is vital to monitor healthcare systems’ resilience, particularly during periods of high distress, and to investigate whether such indicators can be useful to evaluate healthcare systems’ capacity and describe systems’ resilience during emergencies.

Given the forced reorganization of healthcare services during the first wave of the pandemic, we aimed to measure process and outcome indicators related to the quality of care provided to non-COVID-19 elderly patients affected by HF in Emilia-Romagna. Specifically, we analyzed the length of hospital stay (LOS) and timeliness of surgical treatment/rehabilitation as process indicators, and 30-day mortality as the main health outcome.

## MATERIALS AND METHODS

### Setting of the study

The Italian national health service (NHS) is a universalistic health system funded through general taxation. Emilia-Romagna is one of the largest regions of northern Italy, with ∼4.5 million inhabitants as of 2020. Its regional health system includes 8 local health trusts, 4 university hospitals, 1 general hospital trust, and 4 research hospitals.

In 2013, Emilia-Romagna improved the management of patients with HF, reducing the delay of surgery and designing specific modalities for postoperative rehabilitation [7, 25].

### Study design and population

In this retrospective cohort study, we gathered the hospital discharge records (HDRs) of all patients aged ≥65 admitted to the hospitals of Emilia-Romagna with a primary or secondary diagnosis of HF (ICD-9-CM code 820) between January and May 2020 (study period) and between January and May 2019 (control period). We excluded non-residents in Emilia-Romagna, transfers from other hospitals, polytraumas (diagnosis-related group 484–487), diagnoses or history of malignant tumors (ICD-9-CM code 140.0–208.9, 238.6, V10), and cases of COVID-19 using the criteria issued on March 10, 2020 by Italy’s Ministry of Health (ICD-9-CM code V01.82, 079.82, 480.3, V07.0). We excluded patients who died within 1 day of hospital admission without surgery, and those directly admitted to spinal injury units, rehabilitation hospitals or long-term care facilities.

We collected from the health administrative databases of Emilia-Romagna the drug prescriptions of each patient over a lookback period of 3 years to compute the Modified Chronic Disease Score (M-CDS), a drug-based index that has been shown to be a good predictor of 1-year mortality [26].

### Processes of care

Process indicators included LOS, HF surgery (within 2 days, after 2 days or never performed), and rehabilitation within 30 days of hospital admission.

HF surgery was defined as any of the following procedures registered in the HDRs: closed reduction of fracture without internal fixation (ICD-9-CM codes 79.00, 79.05); closed reduction of fracture with internal fixation (79.10, 79.15); open reduction of fracture without internal fixation (79.20, 79.25); open reduction of fracture with internal fixation (79.30, 79.35); total or partial hip replacement (81.51, 81.52).

Through data linkage, we were able to track whether post-acute patients entered bed-based rehabilitation programs in public or private hospital units, community hospitals, or nursing home beds in residential care facilities for the elderly.

### Outcome

The outcome under study was 30-day mortality, i.e., all-cause death within 30 days of hospital admission, either inside or outside the hospital.

### Statistical analysis

Numerical variables were summarized as mean ± standard deviation; categorical variables were summarized as counts (percentages). Comparisons of patient characteristics and process indicators between 2020 and 2019 were investigated with the chi-squared or *t*-test. Differences in 30-day mortality were expressed as odds ratios (ORs) with 95% confidence intervals (CIs), and were adjusted by age, sex and M-CDS via multivariable logistic regression analysis.

All analyses were stratified by month of the year. If a significant difference was present between 2020 and 2019, we further adjusted the analysis by including HF surgery and LOS as additional covariates in the multivariable logistic regression model.

All analyses were performed using SPSS 26.0 (IBM Corp. Released 2019. IBM SPSS Statistics for Windows, Version 26.0. Armonk, NY: IBM Corp) and Stata 15 (StataCorp. 2017. Stata Statistical Software: Release 15. College Station, TX: StataCorp LLC). The significance level was set at 5%; 2019 data did not exhibit any systematic difference with 2017/18 data (data not shown).

## RESULTS

Table 1 shows the characteristics of the 5379 patients with HF included in the study (2531 [47.1%] in Jan-May 2020 and 2848 [52.9%] in Jan-May 2019). Most patients were female (74.2%) and mean age was 84.3 years. No significant differences were found in the distribution of age, sex and M-CDS between 2019 and 2020. However, we observed a significant decrease in hospital admissions for HF in March and April 2020 as compared with the same months of the previous year (March: 530–430 = –100 [–18.9%], *P*-value <0.001; April: 445–598 = –153 [–25.6%], *P*-value <0.001).

**Table 1.**
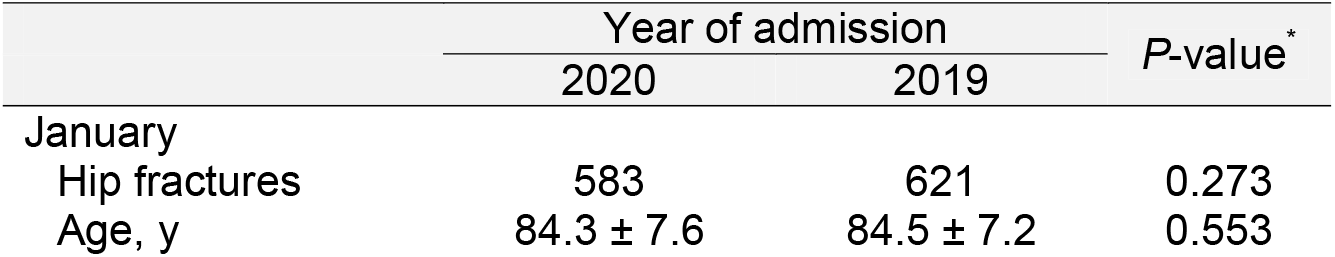
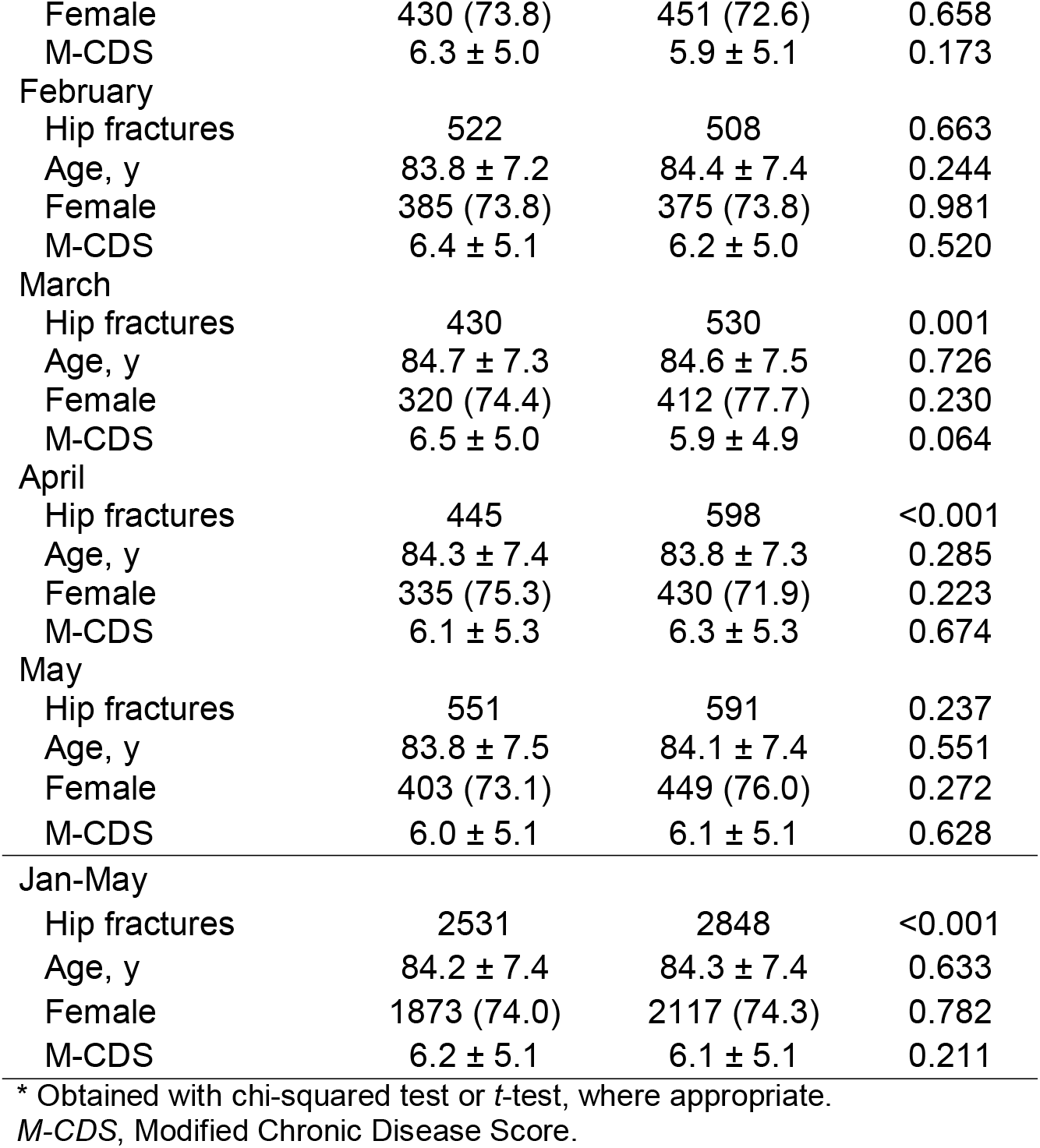
Demographic and clinical characteristics of the study population, by year and month of the year, Emilia-Romagna, Italy. Values are count (percentage) or mean ± standard deviation.

As shown in Table 2, in April and May 2020 there was a significant increase in the proportion of HFs not treated with surgery, as compared with April (10.6–3.5 = +7.1%, *P*-value <0.001) and May 2019 (8.9–4.4 = +4.5%, *P*-value = 0.002), coupled with a significant reduction in the proportion of operations performed within 2 days (April: 70.1–75.6 = –5.5%, *P*-value <0.001; May: 67.3–74.8 = – 7.5%, *P*-value = 0.002). The same data are visually illustrated in Fig 1. Mean LOS was significantly lower in March, April and May 2020 as compared with the same months of the previous year (March: 9.9–11.7 = –1.8 days, *P*-value <0.001; April: 10.9–12.6 = –1.7 days, *P*-value <0.001; May: 11.3–12.3 = –1.0 days, *P*-value = 0.027).

**Table 2.**
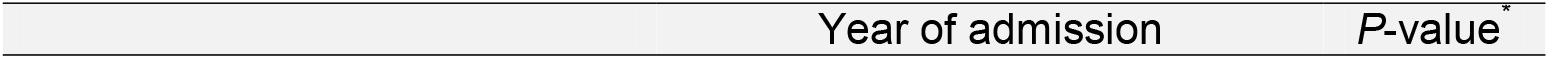
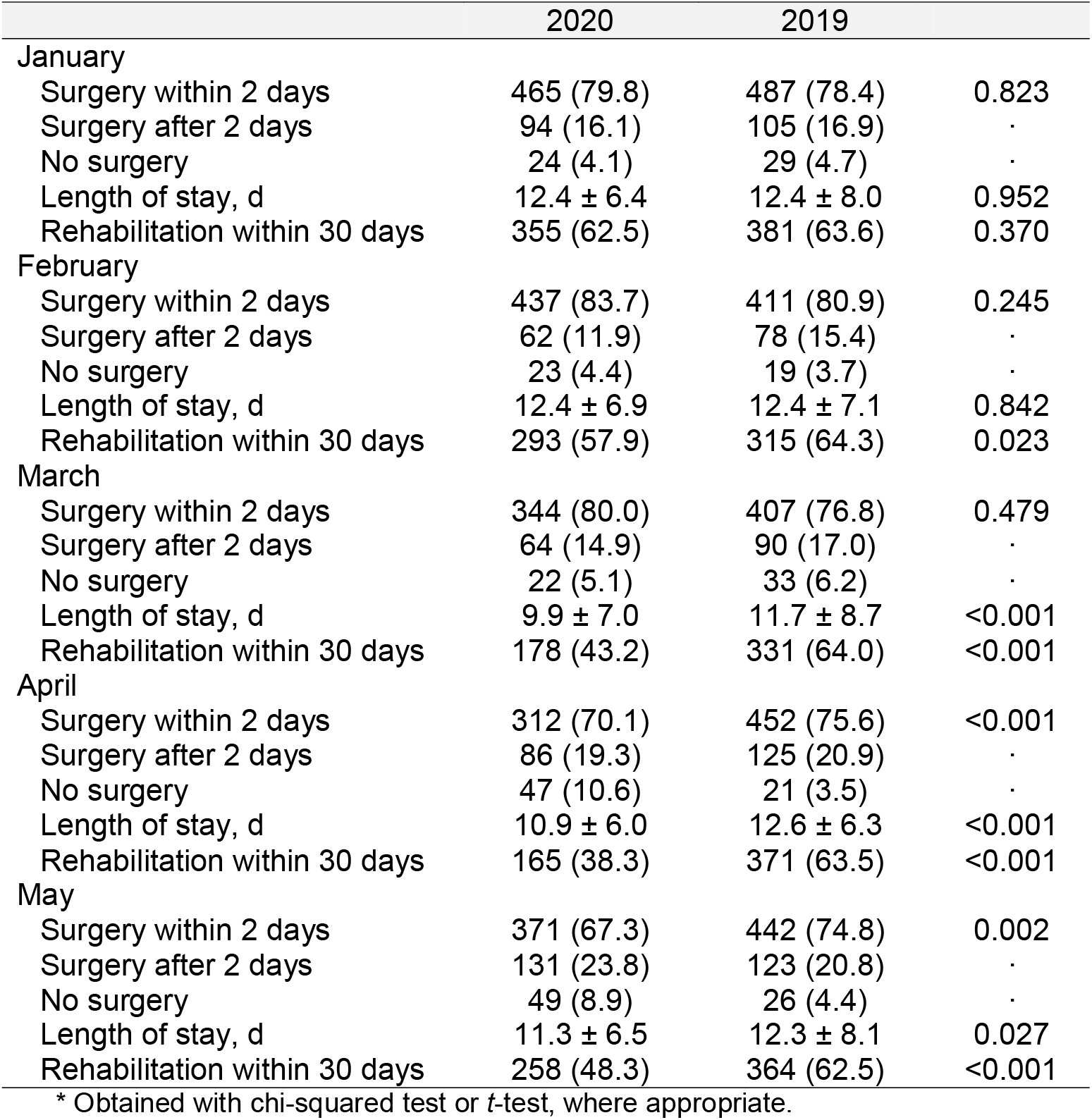
Hip-fracture surgery, length of stay, and rehabilitation by year and month of the year, Emilia-Romagna, Italy. Values are count (percentage) or mean ± standard deviation.

|  | 2020 | 2019 |  |
| --- | --- | --- | --- |
| January |  |  |  |
| Surgery within 2 days | 465 (79.8) | 487 (78.4) | 0.823 |
| Surgery after 2 days | 94 (16.1) | 105 (16.9) | . |
| No surgery | 24 (4.1) | 29 (4.7) | . |
| Length of stay, d | 12.4 ± 6.4 | 12.4 ± 8.0 | 0.952 |
| Rehabilitation within 30 days | 355 (62.5) | 381 (63.6) | 0.370 |
| February |  |  |  |
| Surgery within 2 days | 437 (83.7) | 411 (80.9) | 0.245 |
| Surgery after 2 days | 62 (11.9) | 78 (15.4) | . |
| No surgery | 23 (4.4) | 19 (3.7) | . |
| Length of stay, d | 12.4 ± 6.9 | 12.4 ± 7.1 | 0.842 |
| Rehabilitation within 30 days | 293 (57.9) | 315 (64.3) | 0.023 |
| March |  |  |  |
| Surgery within 2 days | 344 (80.0) | 407 (76.8) | 0.479 |
| Surgery after 2 days | 64 (14.9) | 90 (17.0) | . |
| No surgery | 22 (5.1) | 33 (6.2) | . |
| Length of stay, d | 9.9 ± 7.0 | 11.7 ± 8.7 | <0.001 |
| Rehabilitation within 30 days | 178 (43.2) | 331 (64.0) | <0.001 |
| April |  |  |  |
| Surgery within 2 days | 312 (70.1) | 452 (75.6) | <0.001 |
| Surgery after 2 days | 86 (19.3) | 125 (20.9) | . |
| No surgery | 47 (10.6) | 21 (3.5) | . |
| Length of stay, d | 10.9 ± 6.0 | 12.6 ± 6.3 | <0.001 |
| Rehabilitation within 30 days | 165 (38.3) | 371 (63.5) | <0.001 |
| May |  |  |  |
| Surgery within 2 days | 371 (67.3) | 442 (74.8) | 0.002 |
| Surgery after 2 days | 131 (23.8) | 123 (20.8) | . |
| No surgery | 49 (8.9) | 26 (4.4) | . |
| Length of stay, d | 11.3 ± 6.5 | 12.3 ± 8.1 | 0.027 |
| Rehabilitation within 30 days | 258 (48.3) | 364 (62.5) | <0.001 |

\* Obtained with chi-squared test or *t*-test, where appropriate.

**Fig 1.**
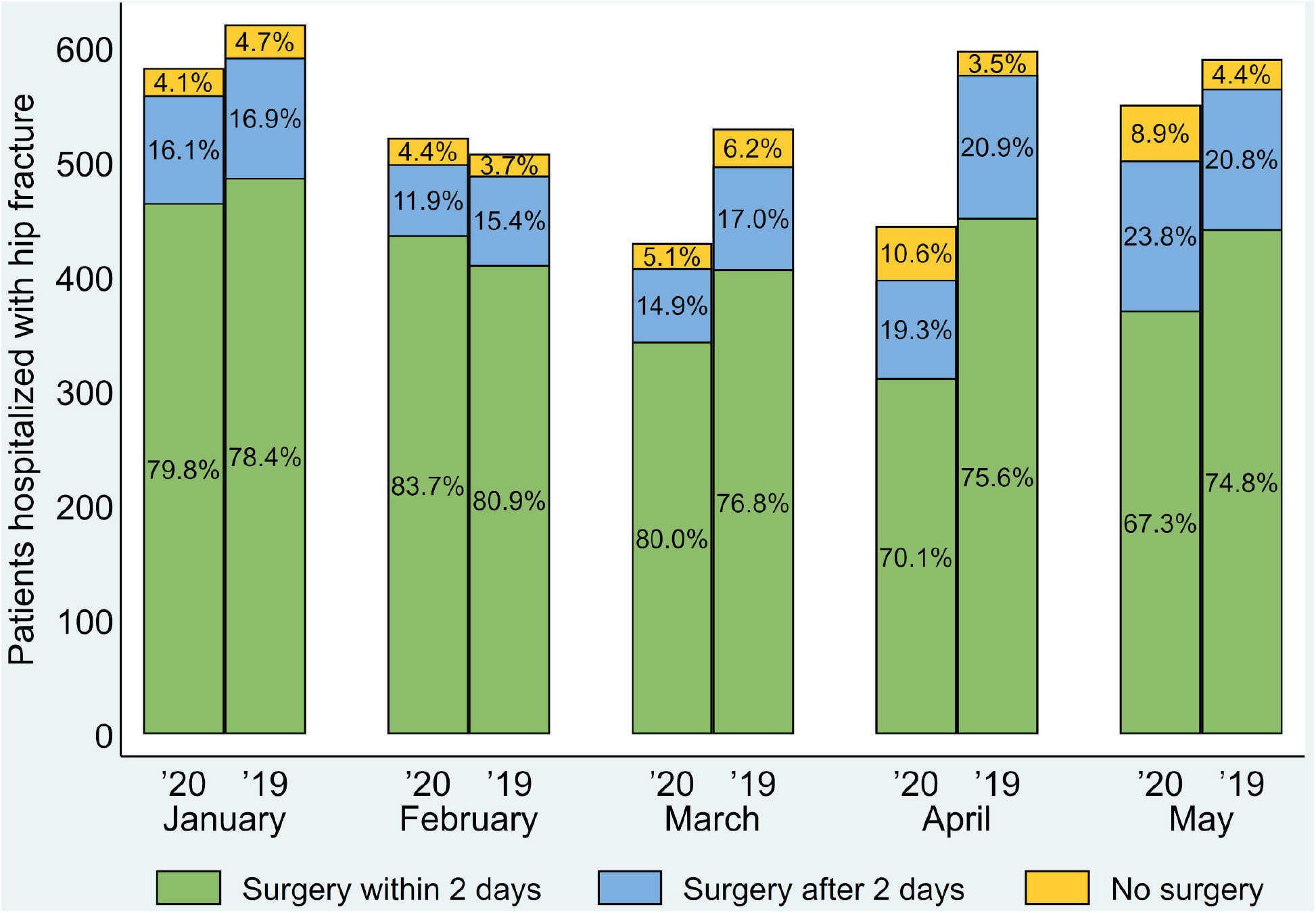
Distribution of hip-fracture surgeries by year and month of the year, Emilia-Romagna, Italy.

In Table 2, we also present the share of HF patients that received rehabilitation treatments within 30 days of hospital admission. We observed a significant reduction as compared with 2019, in particular in February (57.9–64.3 = –6.4%, *P*-value = 0.023), March (43.2–64.0 = –20.8%, *P*-value <0.001), April (38.3–63.5 = –25.2%, *P*-value <0.001) and May (48.3–62.5 = –14.2%, *P*-value <0.001).

Table 3 shows the 30-day mortality rates between January and May, and the corresponding adjusted ORs comparing 2020 and 2019. We found a significant increase in mortality in March 2020 (adj. OR = 2.08, 95% CI = 1.23 to 3. 53, *P*-value = 0.007).

**Table 3.** Thirty-day mortality following hip fracture by year and month of the year, Emilia-Romagna, Italy. Values are count (percentage) unless otherwise specified.

| Month of admission | Year of admission |  | Odds ratio<br>95% (CI) | Adj. <sup>*</sup> odds ratio<br>95% (CI) |
| --- | --- | --- | --- | --- |
|  | 2020 | 2019 |  |  |
| January | 24 (4.1) | 38 (6.1) | 0.66 (0.39–1.11) | 0.67 (0.39–1.15) |
| February | 28 (5.4) | 24 (4.7) | 1.14 (0.65–2.00) | 1.21 (0.68–2.15) |
| March | 41 (9.5) | 24 (4.5) | 2.22** (1.32–3.74) | 2.08** (1.23–3.53) |
| April | 26 (5.8) | 25 (4.2) | 1.42 (0.81–2.50) | 1.48 (0.83–2.64) |
| May | 33 (6.0) | 24 (4.1) | 1.51 (0.88–2.58) | 1.50 (0.86–2.60) |

\*Adjusted by age, sex, and comorbidity index (M-CDS) via logistic regression analysis.
\*\* *P*-value ≤0.01.
CI, confidence interval.

Table 4 summarizes the logistic regression model and shows the association of demographic/clinical characteristics, healthcare process indicators and study period (March 2020 vs. 2019) with 30-day mortality following HF. We found that females had a reduced risk (adj. OR = 0.52, 95% CI = 0.30 to 0.91, *P*-value = 0.023), while patients aged ≥90 had an increased risk as compared with those <80 years (adj. OR = 4.33, 95% CI = 1.70 to 11.04, *P*-value = 0.002). M-CDS and LOS were not associated with increased 30-day mortality, while undergoing surgery after 48 hours since hospital admission and not receiving surgery were significant risk factors (adj. OR = 3.08, 95% CI = 1.59 to 5.97, *P*-value = 0.001; adj. OR = 6.19, 95% CI = 2.86 to 13.38, *P*-value <0.001; respectively). Controlling for these factors, HF patients hospitalized in March 2020 were at higher risk of 30-day mortality as compared with those hospitalized in March 2019 (OR = 2.21, 95% CI = 1.27 to 3.86, *P*-value = 0.005).

**Table 4.** Association of demographic/clinical characteristics, process indicators and study period (pre-/post-pandemic) with 30-day mortality following hip fracture among patients admitted to the hospital in March, Emilia-Romagna, Italy.

| Characteristic | Odds ratio | 95% CI |
| --- | --- | --- |
| Sex |  |  |
| Male | Ref. |  |
| Female | 0.52* | 0.30–0.91 |
| Age, y |  |  |
| <80 | Ref. |  |
| 80–89 | 2.36 | 0.95–5.86 |
| ≥90 | 4.33** | 1.70–11.04 |
| M-CDS |  |  |
| 0–1 | Ref. |  |
| 2–5 | 0.78 | 0.30–2.04 |
| 6–9 | 1.55 | 0.62–3.89 |
| ≥10 | 1.29 | 0.50–3.32 |
| Surgery |  |  |
| Within 2 days | Ref. |  |
| After 2 days | 3.08** | 1.59–5.97 |
| No | 6.19*** | 2.86–13.38 |
| Length of stay, d |  |  |
| <7 | Ref. |  |
| 7–14 | 0.72 | 0.37–1.38 |
| >14 | 0.97 | 0.45–2.10 |
| Study period |  |  |
| Pre-COVID-19 (2019) | Ref. |  |
| Post-COVID-19 (2020) | 2.21** | 1.27–3.86 |

\*\*\* *P*-value ≤0.001, \*\* *P*-value ≤0.01, \* *P*-value ≤0.05.

## DISCUSSION

In this observational study, we assessed the impact of the first wave of the COVID-19 pandemic on non-COVID-19 patients with HF. Specifically, we investigated whether the health crisis affected the quality of care provided to elderly patients by analyzing timeliness of surgical interventions, LOS and share of timely rehabilitation as process indicators, and 30-day mortality as the main health outcome.

### Statement of principal findings

In summary, our study shows that the pandemic negatively affected non-COVID-19 patients with HF. The quality of care has been undermined by the unavoidable services’ reorganization needed to address the emergency. The proportion of patients undergoing surgery and receiving timely treatment decreased, as well as the mean LOS and the timely use of rehabilitation services. Health outcomes suffered as well: patients with HF experienced an increased mortality rate, particularly in March 2020. In the following months, HF mortality returned to pre-crisis levels, demonstrating the adaptation and resilience of the healthcare system.

### Interpretation within the context of the wider literature

Our analysis shows that the first wave of the COVID-19 pandemic determined a significant reduction in HF hospitalizations in Emilia-Romagna, one of the most severely hit areas of Italy and Europe, although the population case-mix did not differ between 2020 and 2019. This finding is consistent with other studies [27-29] and can be explained by the enforcement of a strict national lockdown from March 9 to May 4, 2020. By confining people at home, interrupting mobility and work activities, and reducing road traffic, the frequency of travel- and work-related injuries dropped; this led to an overall reduction in the number of patients accessing emergency departments and hospitals. Fear of hospitalization could also be responsible for this reduction [30, 31].

The disruption of the healthcare services determined an increase in the percentage of patients with HF that did not undergo surgery and a decline in the share of patients undergoing timely surgery within 48 hours of hospital admission, together with a reduction in the mean LOS. These changes could be ascribed to the sudden hospital overload experienced during the first months of 2020, which coerced healthcare institutions to enforce prioritization of their services. Many professionals’ skills, such as surgeons’ and anesthesiologists’, were repurposed to attend to COVID-19 patients in dedicated wards. This created service gaps, reducing both the number of physicians dedicated to non-COVID-19 patients and the time dedicated to each of them [27]. Diminished healthcare capacity was the reason behind the curb of peri- and post-operative care in HF patients, which is shown by the significantly reduced mean LOS. Following the health policy maker’s suggestions, early discharge was recommended to decrease the risk of hospital-acquired infections and to convert non-COVID-19 hospital beds to COVID-19 beds. Other studies described similar gaps in the healthcare services’ capacities and capabilities during the pandemic and reported similar results [28,29].

Further considering health services’ performance, we found that the number of patients receiving bed-based rehabilitation within 30 days of hospital admission from February to May 2020 was lower compared with the same period of the previous year. The performance of rehabilitative care could have been undermined by the difficulty to reorganize treatment pathways for non-COVID-19 patients. In Emilia-Romagna, many rehabilitation centers experienced COVID-19 outbreaks, preventing them from providing adequate standards of care and safety [27]. As shown in several studies, the inability of outpatient rehabilitation facilities (i.e., community hospitals and nursing homes) to accept and treat patients coming from acute care hospitals could be responsible for the reduction and delay of timely rehabilitation treatment [32,33].

Since the initial outbreak of the pandemic, several authors reported an excess of mortality for patients with COVID-19 and affected by HF [34-36]. Our study shows that in 2020 the 30-day mortality rate of non-COVID-19 elderly patients with HF was higher compared with the previous year. This increase was significant in March 2020 (9.5% vs. 4.5%), which was the month with the higher incidence of COVID-19 cases in Emilia-Romagna during our study period (see S1 Fig). The risk of dying (adjusted by age, sex, and comorbidities) was twice as high as the one observed in March 2019. This could be related to the extreme pressure that healthcare structures had to withstand [36], to the abrupt changes in healthcare organization and management, and the possible lack of attention to the treatment pathways [28,29,36].

Multivariable analysis showed that the increase in mortality in March 2020 was associated with relevant processes of care, such as surgical treatment, timely surgery, and LOS. Plenty of literature supports the importance of these factors as predictors of patients’ outcomes, especially mortality [34]. Nonetheless, the variations in the aforementioned process indicators did not fully explain the 30-day mortality difference between March 2020 and March 2019. This important finding reveals the presence of additional factors that could be investigated in further studies. They could be identified in misreporting or misclassification of actual COVID-19 cases, and/or factors related to patient clinical management that we did not evaluate.

During the following months (April and May 2020), we saw an increase in 30-days mortality, albeit not statistically significant. Of note, during these months the process indicators remained significantly worse than in 2019. These findings underline the Emilia-Romagna healthcare system’s capacity to respond to the initial health crisis and to take effective actions to mitigate the impact of the pandemic.

### Implications for policy, practice and research

In this study, we found that the quality of care for patients with HF has been undermined by the unavoidable healthcare services’ reorganization needed to address the COVID-19 pandemic. After the first months of the emergency, HF indicators returned to pre-crisis levels, demonstrating the adaptation and resilience of the healthcare system. However, despite the inability to evaluate functional capacities and medium-/long-term healthcare quality indicators, we can assume that the cumulative unmet needs of the patients that did not receive timely surgery and rehabilitation may lead to a worsening of their medium- and long-term outcomes. In light of this, healthcare policymakers and professionals involved in the management of COVID-19 patients should be aware of the needs of patients with other acute and chronic health needs, which should be carefully considered, investigated, and included in future emergency preparedness and response plans.

### Strengths and limitations

Analysis of complete data related to the whole healthcare system of a wide region is the main strength of this study. Moreover, the Italian experience during the first wave of the COVID-19 pandemic represents a teachable event illustrating the healthcare system early response to a severe health crisis. Misreporting and misclassification of COVID-19 cases and deaths is the main limitation of our study. However, we relied upon the ICD-9-CM classification system issued by Italy’s Ministry of Health and Regional Authorities for the correct identification of COVID-19 cases and deaths. Other limitations are common to all studies based on administrative data, including lack of accuracy and differences in the coding criteria over time, but it is hard to believe that such potential sources of information bias might have significantly affected our estimates.

## Conclusions

This study addressed the impact of the COVID-19-related healthcare reorganization on healthcare quality and 30-day mortality for non-COVID-19 elderly patients with HF. Our results show a reduction in the proportion of patients undergoing surgery and in the share of patients receiving timely surgery and rehabilitation. Mortality increased significantly in March 2020 as compared with March 2019, but differences in patients’ case mix and quality of care only partially explained such increase. Further studies are needed to verify additional determinants, to identify the strengths and weaknesses of healthcare systems and to develop capacities and capabilities suited to face the upcoming public health challenges.

The care and attention required for patients with COVID-19 should not distract from the needs of patients with other critical acute and chronic conditions, which should be carefully investigated and included in future emergency preparedness and response plans.

## Data Availability

The data that support the findings of this study are available from Emilia Romagna Regional Healthcare Information System, but restrictions apply to the availability of these data, which were used under license for the current study, and so are not publicly available.

## Contributorship

DG and FS had the idea, contributed to study design and data interpretation, and drafted the manuscript; AC, FE and GG contributed to design, data acquisition and interpretation, and drafted the manuscript; FS contributed to design and data acquisition, and drafted the manuscript; SR contributed to data acquisition and analysis; MA contributed to data acquisition and interpretation; MPF and JL contributed to conception and data interpretation, and critically revised the manuscript. FS and SR have verified the underlying data.

All authors gave their final approval and agreed to be accountable for all aspects of the work.

## Ethics and other permissions

This study was approved by the Comitato Etico Indipendente di Area Vasta Emilia Centro (approval: April 17, 2019; amendment: March 22, 2021). Data used in this research were obtained from the Regional Healthcare Information System, which includes detailed information on the use of healthcare services by all regional patients, with the patient as our unit of observation. The study, based on routine administrative information, was carried out in conformity with the regulations on data management of Emilia-Romagna and with Italian privacy law.

## Supporting information Caption

**S1 Fig. Incidence and prevalence of COVID-19 cases (×100,000 population) in Emilia-Romagna, Italy, between February 24, 2020 and May 31, 2020.** Source: *Dipartimento della protezione civile*.

