## Supplement for "Variations of the quality of care during the COVID-19 pandemic affected the mortality rate of non-COVID patients with hip fracture"

**Supporting information**

**S1 Fig. Incidence and prevalence of COVID-19 cases (×100,000 population) in Emilia-Romagna, Italy, between February 24, 2020 and May 31, 2020.** Source: *Dipartimento della protezione civile*.

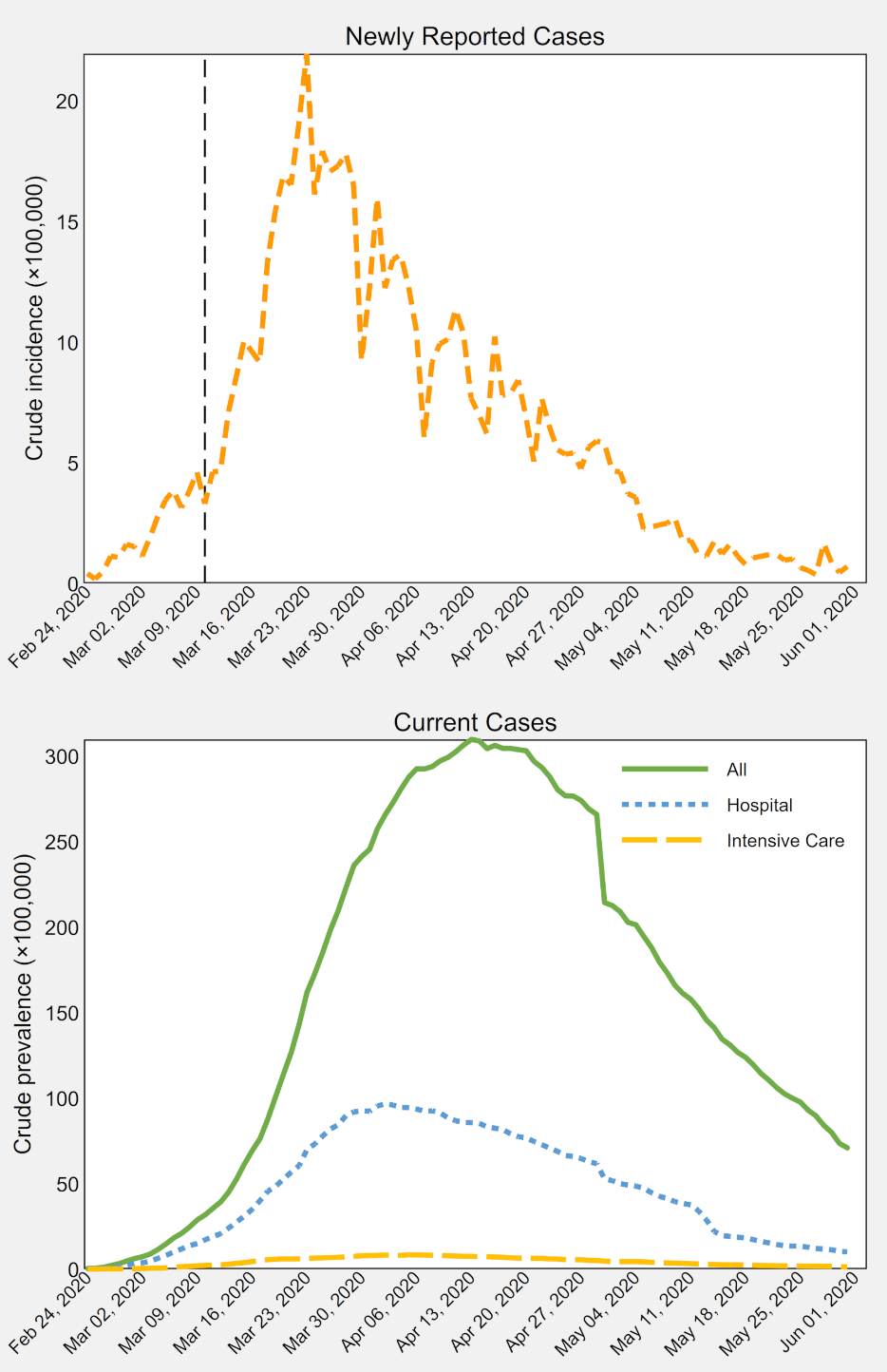
